# PERSONAL PROTECTIVE EQUIPMENT (PPE) USING IN ANTALYA 112 EMERGENCY AMBULANCE SERVICES DURING OUTBREAK

**DOI:** 10.1101/2020.06.16.20129171

**Authors:** Mehmet Fatih Gulsen, Münevver Kurt, Isil Kaleli, Ayhan Ulasti

**Affiliations:** Directorate of Antalya Emergency Medical Service; Gerontology, Faculty of Health Sciences, Akdeniz University (PhD student)

**Keywords:** COVID 19, Ambulance Services, PPE, Personal Protective Equipment

## Abstract

A new type of Coronavirus (SARS-CoV-2) was identified on January 7th, 2020, and the disease was named COVID-19 caused by this virus. The first confirmed COVID-19 (+) case in Antalya was detected on March 14^th^, 2020 and transferred by an Antalya 112 emergency ambulance to hospital.

Using of Personal Protective Equipment (PPE) in ambulance work standards against COVID-19, was recommended by global health authorities. The aim of this descriptive, retrospective and cross-sectional study which is conducted between the dates of March 14th, 2020 – May 31st, 2020, is to evaluate the level of PPE usage, the risk analysis results of Antalya 112 ambulance teams during outbreak. There were 5344 possible and 787 confirmed COVID-19 (+) positive ambulance cases between the dates of study conducted. The majority of these cases were male (62%) and over the age of 65 (47%). The majority of cases were result in “transferred to healthcare facilities” (75.48%). The total 2361 ambulance team workers were taken to risk analysis measurements in terms of COVID-19. The majority of ambulance team members were assessed with no risk available. The ambulance team members who are assessed with no risk available, has preferred to use Level-4 PPE (N95/FPP-Goggle/face protection-Gloves-Apron/coverall) mostly (84.50%) during ambulance transfers. Using mask on patient and the level of PPE usage showed negative correlation with risk level of Healthcare Workers (HCWs). There have been detected only 2 HCWs diagnosed with COVID-19 with Computed Tomography. The follow-up procedures of two HCWs has been finalized, and both of them cured. The studies about easy-to-use, high-tech PPE with maximum protection, are recommended for further studies.

## 1. INTRODUCTION

The World has been witnessing of various pandemics in the last two thousand years. The plague of Justinianus (AD542-546), the Bubonic Plague (1347-1351), also known as Black Death, and Smallpox (1520) are among the most life-threatening outbreaks (1). Influenza outbreaks (2) caused by Influenza A virus, have been occurred three times only in the 21st century. The most severe is “Spanish Flu” (type A H1N1), which was estimated to have killed more than 50 million people in 1918-1919 and others Asian Flu (type A H2N2) in 1957-1958 and the Hong-Kong flu (type A H3N2) in 1968-1969. During the H1N1 pandemic that emerged in 2009, it is thought that there were more than 622,000 confirmed cases and at least 12,220 deaths in more than 208 countries at the end of the same year (3).

The coronaviruses (CoVs) were first isolated in 1965 by Tyrrell and Bynoe in the culture taken from the human respiration channel. Coronaviruses, that with single chain, positive polarity, enveloped RNA and frequently mutated, also affect the animals and they are the reasons of the 15% flu cases in human society. These viruses are divided into four types according to current taxonomy: alphacoronavirus, betacoronavirus (SARS-CoV and MERS-CoV), gammakoronavirus and deltacoronavirus (4). MERS-CoV was first described in humans in Saudi Arabia, September 2012. SARS-CoV had previously emerged as an international health emergency that caused hundreds of people to die in 2003. Finally, a new type of Coronavirus (2019-nCoV) was identified on January 7^th^, 2020, after the WHO (World Health Organization) China Office reported unknown the cases on December 31^st^, 2019, in the city of Hubei, Wuhan, China. The new diseases were entitled as COVID-19 and the agent virus was named SARS-CoV-2 because of its similarity to previous SARS CoV (5). The first imported case a 61-year-old Chinese woman in Thailand was reported on January 13^th^, 2020. The first COVID-19 case in Turkey was detected on March 11^th^, 2020 (6). The first COVID-19 patient who transported by 112 emergency ambulance on March 14th, 2020, was first case of Antalya.

### 1.1. Managing Human and Material Resources In Ambulance Services During Outbreak

The fight against pandemics (outbreaks) should be managed with international and intersectoral aspect. Ethical and legal issues are needed to be considered during preparing and managing of outbreaks. While adapting to national and international decisions, regional plans should be carried out by prioritizing access to healthcare resources during implementation of these plans. There are many ethical problems and difficult questions to be answered, which should be taken account by authorities (7). In order to stop the COVID-19 outbreak turning into a global health emergency situation, the governments have had to carry out their epidemic control activities with many uncertainties under the increased pressure (8).

The news about ambulance and staff insufficiency have come out (9, 10) during outbreak in press. Ambulance teams have struggled to find health facilities to deliver the patients waiting in ambulance queues in front of the hospitals (11, 12). It has been stated that the use of public transport or personal vehicles to access hospitals, increases the transmission risk. Therefore, preliminary evaluation at home and avoiding hospital application without suspicious and severe symptoms have been recommended (13). In order to prevent risks, reduce the unnecessary use of hospitals and ambulances, the home testing method have been applied in London, in cooperation of ambulance service providers (14). The 90% percentage of test in the city was perfomed in homes (15). But long-term benefits of home testing method on human and material resources are unknown whether stop COVID-19 outbreak. Moreover “Hospital transfer” is still needed for further examination and treatment.

WHO (16) recommends the use of gloves, masks and eye protection in minimum, and also emphasizes to adequate PPE (Personal Protective Equipment) supply to meet minimum PPE requirement, proper PPE use and PPE optimization. Guideline of the rational use of PPE is also published in case of shortage of PPE (17). The Centers for Disease Control and Prevention (CDC) has specified the standards of PPE use in emergency calls and during transfers according to the job description of healthcare workers (HCWs), distance to suspected patient and procedures performing in ambulances (18). In the pre-hospital emergency health care service recommendation guide published by WHO’s Pan-American Health Organization (19), type of PPE use varies according to the interventions made in ambulance like aerosol generating procedures (AGPs), job description of staff, distance to patients, condition of patient. In Ambulance Safety Guide (20) prepared by UK Public Health Authority, the use of PPE is strongly recommended to prevent and control COVID-19 outbreak.

### 1.2. Global Strategies against COVID-19 for Ambulance Services

The importance of prevention and control strategies, has been realized to protect HCWs against COVID-19 in many country (21, 22). WHO Pan American Health Organization and UK Public Health Authority gives important recommendations about prehospital emergency service process with administrative and operational approaches which start from call management till end of the patient transport included PPE use standards and ambulance decontamination during COVID-19 outbreak (19, 23).

Since demands of ambulance services extraordinarily increased while rising ambulance use hesitations due to high risk of transmission in the narrow area; the studies related rapid and safe ambulance disinfection and minimum PPE use (24, 25, 26) accelerated. The validity of the PPE standards in the transfer of aircraft cabin which is rather limited area, has been criticized (27) and become the subject of debate. How should be use of PPE in emergency ambulances with narrower area than aircraft cabin? What should be the minimum PPE to be used by the ambulance team who travel 20-30 minutes average with the patient in a narrow space during outbreaks?

CDC (28) has published guidelines about operations of emergency services and 911 call centres by particularly emphasizing importance of supply and use of PPE in ambulances during outbreak. General isolation measures and individual protection methods are also prepared and presented in COVID-19 guide in China where the first case reported (29).

In the interview dealt with three Canadian experts, consensus have been provided about priority allocation of PPE (especially N95 masks) to front-line healthcare professionals by eliminating access difficulty (30). Although there are several difficulties reported by HCWs related PPE use, they have the competence and willingness to comply with infection prevention and control guidelines. Participation of HCWs to preparation of guides by allowing the inclusion of their decisions, will facilitate to follow instructions well (31). According to the results of a study conducted in Singapore, well-organized staff protection system which is implemented in work routines is quite effective to protect frontline HCWs, if it is implemented in non-outbreak period (32).

### 1.3. National Strategies against COVID-19 for Ambulance Services

Before the COVID-19 outbreak, required regulations and arrangements were already prepared for the implementation of Turkish ambulance services in case of outbreak. According to the Turkish Infectious Diseases Surveillance and Control Principles Regulation (33), epidemiological surveillance, inter-institutional data sharing, an early warning and response system establishment, disease-specific surveillance and control improvement studies and interventions are required to prevent and control of communicable diseases. The measures to be taken against infections during patient transportation and ambulance cleaning, are notified in the guide prepared by Authorities of Ministry of Health. The guide emphasizes the protection of HCWs and the use of PPE correctly (34).

Preparedness for the outbreak had been already started before the first COVID-19 case was detected in Turkey. The scientific committee and 7/24 teams were formed; instructions, guides, algorithms, and documents were prepared and delivered to end users by electronically and physically through the communication network created according to National Pandemic Plan of Turkey (35). General Directorate of Public Health have prepared a website for infection control studies to manage COVID-19 outbreak (36) and conducted guidebook which is updated as needed (6). All of these studies have guided the establishment of PPE standards for ambulance-use during outbreak.

These standards suggest different type of PPE using, depending on whether the patient is suspected, the job description of HCWs (paramedic, ambulance driver, or cleaning staff), exposure level to patient, type of procedures operated in ambulance (37). The visual guidance tools have been prepared about the minimum PPE requirements, practical instructions of gloves, masks, eye and face protectors and aprons for healthcare professionals (38).

### 1.4. Strategies of Antalya Ambulance Services against COVID-19

Emergency call strategies during COVID-19 outbreak, vary from country to country according to allocation policies of HCWs, ambulances, drugs, materials and equipment (39, 40, 41, 42). Antalya Ambulance Services Directorate is responsible authority for 61 Emergency Medical Services (EMS) Stations which give healthcare services with 101 ambulances (included one helicopter ambulance) and 972 HCWs. The EMS ambulance stations are directed to cases according to the calls received from 112 Call Centre; calls and operation procedures are managed according to the standard algorithms in routine.

COVID-19 Emergency Response Team has been established within the Antalya Ambulance Services Directorate coordinated with Crisis Coordination Centre of Antalya Provincial Directorate of Health. All procedures during outbreak related with possible / confirmed cases in prehospital process, are managed by this team. This team has developed applications, algorithms, staff trainings for 112 call centre and ambulance stations during outbreak. The strategies of emergency calls and case management are updated when necessary by this team. Some of the implementations during outbreak in Antalya EMS stations:

1. Work schedule planning team is formed, and problem-solving strategies and human resources with alternative scenarios created in case of over demand.
2. Ambulance equipment, device and material tracking planning team is formed, supply strategies and distribution chain with alternative scenarios created in case of over demand.
3. The ULV (ultra-low-dose) ambulance disinfection method is applied for 6 minutes by using hydrogen peroxide and colloidal silver. After each UVL procedure, the ambulance door are closed for four minutes and then cabin is ventilated for 15 minutes.
4. Work Health and Safety Committee carry out planned visits to EMS ambulance stations by informing HCWs about health and work safety issues to protect themselves.
5. PPE store and usage status is determined; supply strategies and distribution chain created.
6. A theoretical and practice staff training program is established; PPE reports discussed with teams; feedbacks got from the teams; PPE related procedures updated if needed.
7. PPE usage reports are analysed, and standards of PPE use are updated according to global practices if needed.

In the joint mission report of the WHO-China on COVID-19, it was recommended to use negative pressure ambulances and conduct studies subjected to the most effective PPE usage in practice (43). Since the routine use of PPE in work standard requires extra time and effort, many difficulties come out in terms of optimum and accurate PPE usage additional to material shortage. In this study we aimed to evaluate the use of PPE, by comparing the number of COVID-19 involved cases / infected HCWs.

## 2. METHOD

The Most of phone calls which is addressed to 112 Call Centre of Antalya are named as a “case” except the unfounded notice, wrong number, case-reject etc. After call centre staff takes information about cases, one of the ambulance team (among 61 Antalya Ambulance stations) is appointed for each case according to location of patient.

This study is initial attempt to identify optimum PPE requirements to be used at emergency ambulance services for future outbreaks. It is aimed to evaluate the PPE use in Antalya 112 ambulance services by comparing risk level of HCWs in this study. This descriptive, retrospective, cross-sectional study has been conducted between the dates of March 14th, 2020 – May 31st, 2020.

### 2.1. Triage Procedure in Antalya Emergency Services

The possible/confirmed COVID-19 cases are first eliminated by the 112 Call Centres according to COVID-19 triage questions which are indicated in the guide prepared by Scientific Committee, Ministry of Health of Turkey. In case of the answer of the triage questions are “yes” for more than 2 in 5 questions, the patient evaluated risky in terms of possible COVID-19 infection. Triage questions of 112 call centre (6):

1. Do you have a cough?
2. Do you have dyspnoea or breathing difficulties?
3. Do you have a fever, or did you have a fever recently?
4. Has any of your relatives been hospitalized for respiratory disease in last 14 days?
5. Has any of your relatives diagnosed with COVID-19 in last 14 days?

The ambulance staff are directed to the use of gloves, medical masks and goggles/face protection as minimum for ordinary cases. For suspected cases, it is recommended that ambulance team has to use gloves, N95 / FFP2 mask, and goggles / face protection in minimum. No companion is accepted to ambulance for adult patients. In case of absolute necessity-like parents of paediatric patients needed-companions are accepted in cabin with a surgical mask (6).

### 2.2. Risk Assessment of Ambulance Teams in Antalya Emergency Services

Every EMS station with 7/24 team is on duty consisted of 3 HCWs (driver, 2 paramedics and/or emergency medicine technician) minimum. Eight of the 61 EMS station give services with teams including medical doctor. Daily team managers are appointed for each shifts. After teams finalize patient transfers, possible COVID-19 cases are confirmed in healthcare facilities in 24 hours by using not only PCR (polymerase chain reaction) but also CT (Computed Tomography) scan for more accurate results by the reason of which is shown in studies conducted (44, 45, 46). Healthcare facility notifies case information (date, time, name of ambulance teams) about COVID-19 (+) ambulance patients to COVID-19 Emergency Response Team (see section 1.2.2) in 24 hours. The Emergency Response Team determines the level of PPE usage not only by checking ASOS records, but also by asking team managers of ambulances. Team manager is responsible to report PPE usage of each team member. The team is assessed in terms of risk level of exposure and level of PPE usage by using the algorithm named “Risk Level Assessment of HCWs who exposed to COVID-19” (As shown Table 1) (47).

**Table 1:** Risk Level Assessment of HCWs who exposed to COVID-19

|  | **PPE Usage of HCWs** | **Exposure risk** |
| --- | --- | --- |
| Intensive exposure with patient who wear medical (surgical) mask | No use of medical mask or N95 / FFP2, or Medical mask usage instead of N95 / FFP2 | Moderate |
|  | No use of eye protection | Low |
|  | No use of gloves and apron | Low |
|  | Proper use of all PPE | No risk assessment |
| Intensive exposure with patient who does not wear medical (surgical) mask | No use of medical mask or N95 / FFP2 | High |
|  | Medical mask usage instead of N95 / FFP2 | Moderate |
|  | No use of eye protection | Moderate |
|  | No use of gloves and apron | Low |
|  | Proper use of all PPE | No risk assessment |

### 2.3. PPE Use of Antalya Ambulance Service Stations

The minimum PPE usage procedures are quite similar in Antalya ambulances compare the global applications. The using procedures and recommendation of PPE are stated in the COVID-19 guide (6) prepared by Scientific Committee of Health Ministry of Turkey. After the Call Centre triage in terms of COVID-19 risk, following PPE instructions applied in Antalya Ambulances for all cases:

✓ During PPE wearing, apron/coverall, mask, glasses/ face protector, gloves in order, S During PPE removal, gloves, glasses/ face protector, apron/coverall order in order, S The removal order of glasses/ face protector and apron/coverall may be changed in case whole contamination of apron/ coverall with gloves together,S The mask is removed after patient delivery, gloves out and hand hygiene performed,
✓ After patient delivery to health institution, gloves changed, hand hygiene performed, ambulance cleaning done,
✓ Minimum N95/FFP2 mask – goggles / face protection – gloves and apron used in ambulance cleaning; extra PPE added if needed,
✓ Additional PPE (double gloves, apron, overalls, N95) are used if needed,
✓ Hand disinfection recommended between wearing and removing pieces.

The level of PPE use grading into 4 levels by the aim of easy risk analysis and follow-up procedures of HCWs. But the HCWs are allowed to add extra PPE when needed:

Level-1: Medical mask – Gloves

Level-2: Medical mask – Goggles / face protection – Gloves

Level-3: N95/FFP2 – Goggles / face protection – Gloves

Level-4: N95/FPP – Goggles / face protection – Gloves – Apron / coverall

The Emergency Response Team evaluates the level of risk analysis results by asking team managers and then compares with ASOS records (Emergency Health Information Systems). ASOS is the ongoing system included records of patient information (diagnose, interventions, treatments), materials and equipment used in ambulances are available in and ambulance team members make case records by using their own passwords. Thus, teams declare the type, number and level of PPE usage during intervention of each case. The team manager has to enter code of materials and equipment used in ambulance to control and follow warehouse of EMSs.

### 2.4. Statistical analysis

In this study, COVID-19 (+) confirmed ambulance case information, level of PPE usage of ambulance teams, and risk analysis results of HCWs, were statistically analysed. The data about patients, PPE usage for each patient and for each HCWs, are provided from ASOS records. PPE usage are compared to the level of risk analysis results measured by the Emergency Response Team. The results of the risk analysis of HCWs and characteristics of confirmed COVID-19 (+) cases were analysed with frequency and percentage tests according to months. The using medical mask on patient and the level of PPE used by HCWs were compared with the level of risk assessments by using Pearson Chi-square test. SPSS Statistics (Base v23) program was used to analyse to data.

## 3. FINDINGS AND DISCUSSION

The 115,377 phone calls in total, were assessed by call triage and 5344 of them evaluated as COVID-19 possible cases intervened by Antalya EMS Ambulances between the dates of March 11th, 2020 – May 31st, 2020.

The 787 COVID-19 (+) confirmed cases were reported to Directorate of Antalya Ambulance Services by healthcare facilities within 24 hours after case intervention finished. The most of confirmed cases were diagnosed by CT scan (519; 66%) and 268 (34%) of them by PCR test. The distribution of diagnose method of confirmed cases are shown by month (As shown Table 2).

**Table 2:**
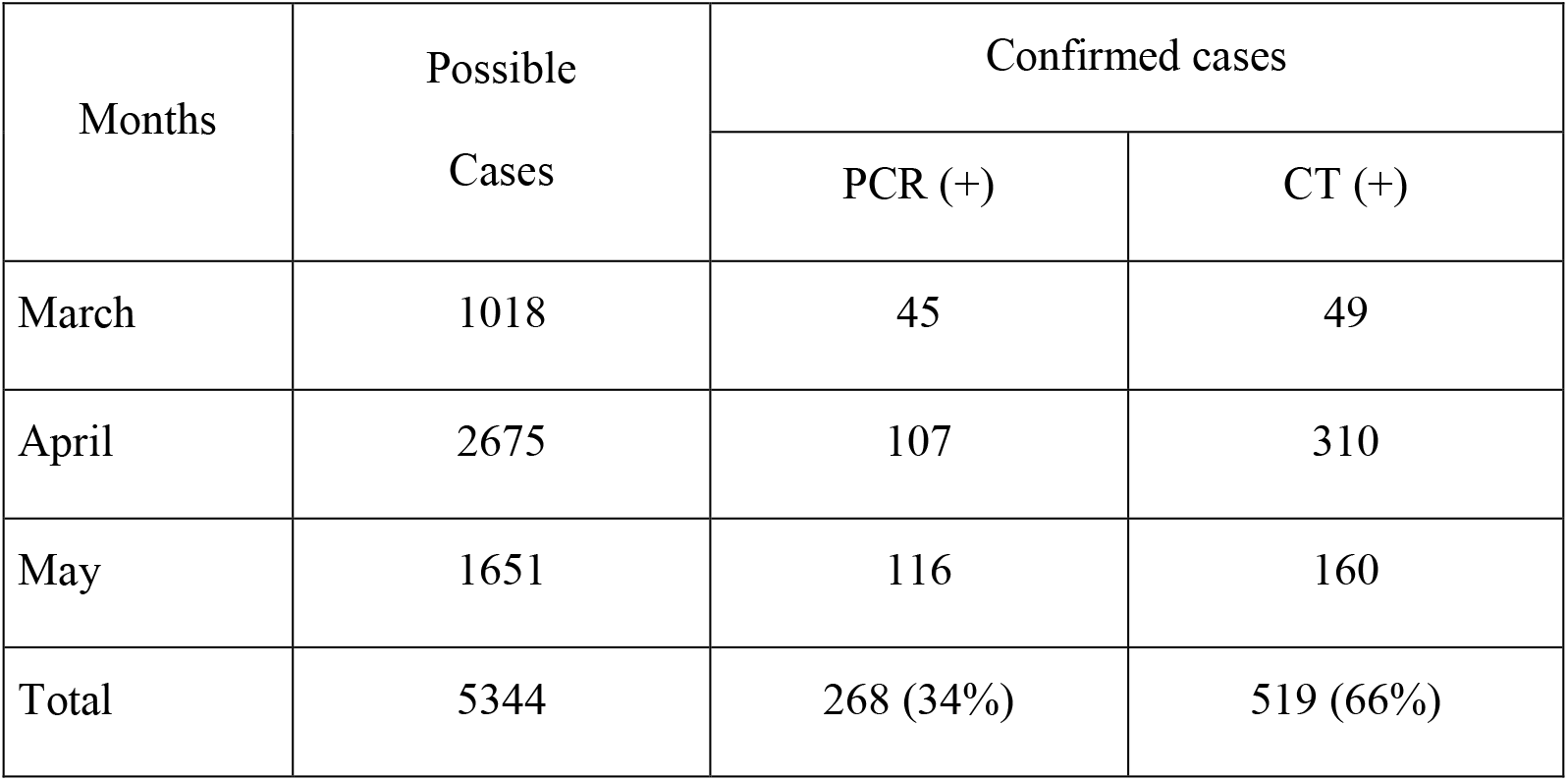
Distribution of Possible and Confirmed COVID-19 Cases by Months (N: 787)

According to age distribution of COVID-19 (+) confirmed ambulance cases, the 65+ patients are the most common group (372; 47%) and the group of 51-64 ages (140; 18%) following (As shown Table 3).

**Table 3:** Distribution of Confirmed COVID-19 Cases by Age Groups (N: 787)

| Months | PCR (+) |  |  | CT (+) |  |  | Total |  |
| --- | --- | --- | --- | --- | --- | --- | --- | --- |
|  | March | April | May | March | April | May | N | % |
| 0-10 | 0 | 1 | 15 | 0 | 3 | 4 | 23 | 2.92 |
| 11-20 | 3 | 5 | 17 | 1 | 5 | 5 | 36 | 4.57 |
| 21-30 | 2 | 19 | 24 | 0 | 9 | 5 | 59 | 7.50 |
| 31-40 | 2 | 24 | 7 | 4 | 22 | 7 | 66 | 8.39 |
| 41-50 | 5 | 17 | 14 | 3 | 40 | 12 | 91 | 11.56 |
| 51-64 | 7 | 15 | 20 | 7 | 59 | 32 | 140 | 17.79 |
| 65+ | 26 | 26 | 19 | 34 | 172 | 95 | 372 | 47.27 |
| Total | 45 | 107 | 116 | 49 | 310 | 160 | 787 | 100.00 |

Increased risk of COVID-19 infection for older adults with a range of other chronic underlying conditions (48) may be a reason for the highest percentage of confirmed cases of 65 and over age. The curfew imposed on the people over aged 65, could be another reason for this people to use 112 ambulances to access to hospital in safe and fast. Gender of the confirmed patients are mostly men (486; 62%) (As shown Table 4).

**Table 4:** Distribution of Confirmed COVID-19 Cases by Gender (N: 787)

|  |  | March | April | May | N (%) |
| --- | --- | --- | --- | --- | --- |
| Men | PCR (+) | 24 | 76 | 60 | 486 (61.75) |
|  | CT (+) | 27 | 197 | 102 |  |
| Women | PCR (+) | 21 | 32 | 56 | 301 (38.25) |
|  | CT (+) | 22 | 112 | 58 |  |
| Total |  | 94 | 417 | 276 | 787 (100.00) |

When team of ambulance departed for a case duty, the results of case could be differed according to general condition of patient and/or decision of teams. The highest percentage (75%) of confirmed COVID-19 cases, resulted in transferring to healthcare facilities. Please rest of the case results as shown Table 5. The rate of “transfers to healthcare facilities” is the most common result of ambulance cases at non-outbreak period as well. The “Transfer to Quarantine place” was emerged as a new type of result for ambulance cases during outbreak.

**Table 5:** Distribution of Confirmed COVID-19 Cases by Case Results (N: 787)

|  | PCR (+) |  |  | CT (+) |  |  | Total |  |
| --- | --- | --- | --- | --- | --- | --- | --- | --- |
|  | March | April | May | March | April | May | N | % |
| Examination in place | 0 | 0 | 0 | 1 | 0 | 2 | 3 | 0.38 |
| Transfer to a healthcare facility | 36 | 69 | 93 | 36 | 239 | 121 | 594 | 75.48 |
| Transfer between facilities | 8 | 13 | 3 | 6 | 46 | 27 | 103 | 13.09 |
| Transfer to Quarantine place | 0 | 22 | 17 | 1 | 5 | 1 | 46 | 5.84 |
| Transfer refuse | 1 | 2 | 3 | 5 | 16 | 8 | 35 | 4.45 |
| Transfer to home | 0 | 0 | 0 | 0 | 1 | 1 | 2 | 0.25 |
| Exitus, left in place | 0 | 1 | 0 | 0 | 3 |  | 4 | 0.51 |
| Total | 45 | 107 | 116 | 49 | 310 | 160 | 787 | 100.00 |

The percentage of “transfer refuse” was 4.5% in confirmed cases. But in the whole cases (ordinary + possible cases: 115,777), “transfer refuse” was higher (March: 14%; April: 17%; May 14%) during outbreak than non-outbreak times that 11 % average. This means that if patients do not suffer from symptoms, they do not prefer to use an ambulance to access healthcare services. Fear of exposed to COVID-19 during transfer or in hospital, could be another reason for higher rate of ambulance transfer rejection.

According to the results of “Risk Level Assessment of HCWs who exposed to COVID-19” algorithm, the use of PPE is evaluated (As shown Table 6). For 787 case (patient), 2361 HCWs in total were on ambulance duty. According to profession, 73 of them are doctor, rest of them paramedic or emergency medicine technician.

**Table 6:**
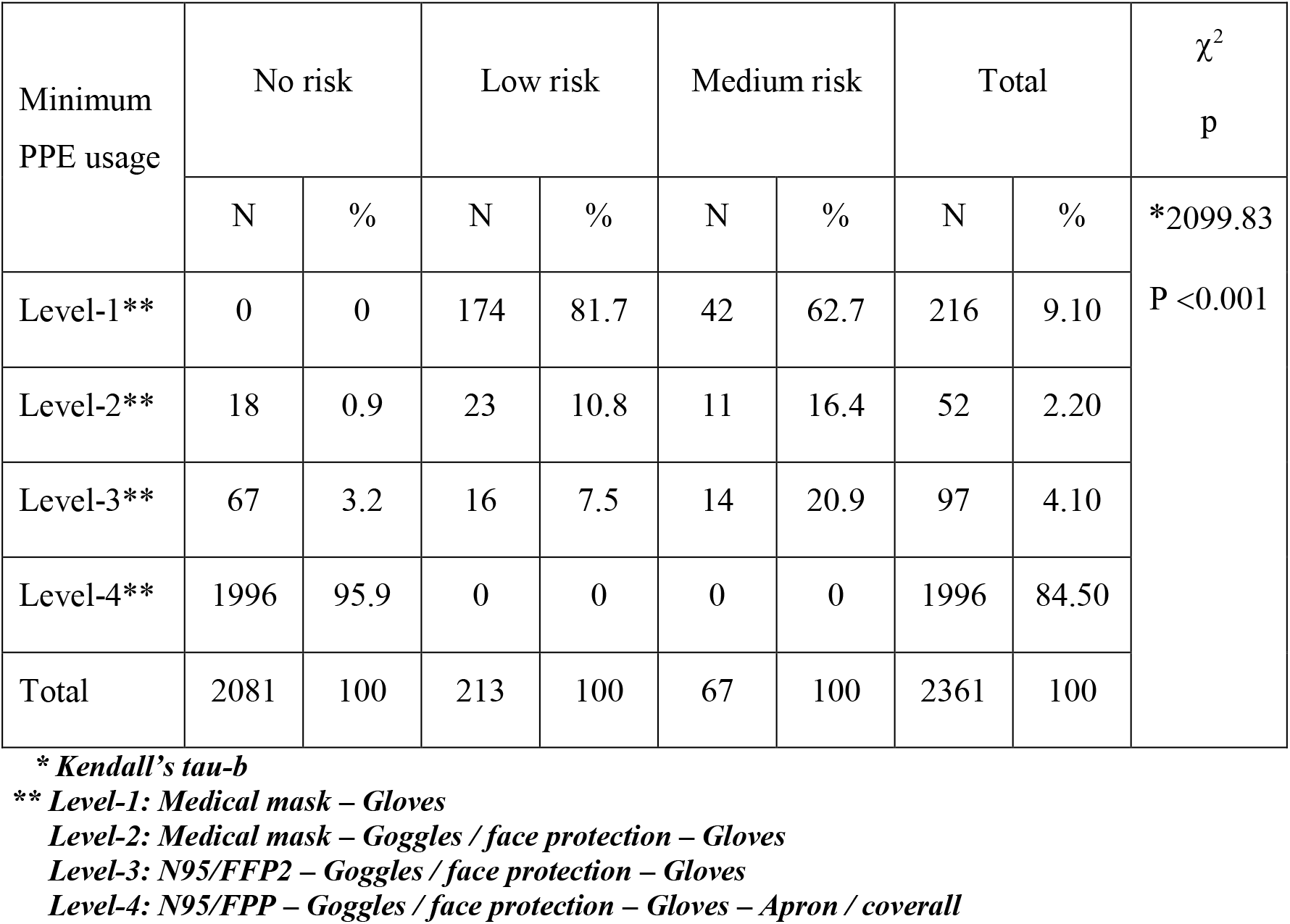
The PPE Usage of HCWs by Risk Level Analysis Results (N: 2361)

| Minimum PPE usage | No risk | | Low risk | | Medium risk | | Total | | $\chi^2$<br>p |
| --- | --- | --- | --- | --- | --- | --- | --- | --- | --- |
|  | N | % | N | % | N | % | N | % | *2099.83<br>P <0.001 |
| Level-1** | 0 | 0 | 174 | 81.7 | 42 | 62.7 | 216 | 9.10 |  |
| Level-2** | 18 | 0.9 | 23 | 10.8 | 11 | 16.4 | 52 | 2.20 |  |
| Level-3** | 67 | 3.2 | 16 | 7.5 | 14 | 20.9 | 97 | 4.10 |  |
| Level-4** | 1996 | 95.9 | 0 | 0 | 0 | 0 | 1996 | 84.50 |  |
| Total | 2081 | 100 | 213 | 100 | 67 | 100 | 2361 | 100 |  |

\* *Kendall's tau-b*
\*\* *Level-1: Medical mask – Gloves*
*Level-2: Medical mask – Goggles / face protection – Gloves*
*Level-3: N95/FFP2 – Goggles / face protection – Gloves*
*Level-4: N95/FFP – Goggles / face protection – Gloves – Apron / coverall*

The Level-4 PPE mostly used for confirmed cases although PPE Level-3 recommended as general standard for possible cases. The Level-4 PPE usage is getting increase when risk level decrease. There is strong negative relation between the level of PPE use and risk level of HCWs (p < 0.001). If the teams use Level-4 PPE, risk level of HCWs evaluated with no risk available (1996; 96% in no risk; 84.5% in total). Because emergencies are unexpected cases may come up suddenly with risky intervention indications like AGPs, intubation indications which are needed to unmask patient, ambulance teams have been ready for worst situation during transfers. Thus ambulance team have added extra gloves, coverall and sometimes aprons to routine PPE standard as a precaution. The total number of HCWs taken to risk analysis and then followed-up procedures were 2361; and 67 of them identified under the medium risk and 2081 of them under no risk assessment. In addition to the standard PPE usage of the ambulance teams, the number of extra added PPE, could not be examined due time restriction and the lack of records and statistical data. Because there is real-time observation technology available in ambulances full-time PPE usage of HCWs could not be detected. PPE which applied at the beginning of case start, could be use incorrectly or taken off sometimes during patient transfer due to emergency intervention stress.

The distribution of confirmed COVID-19 (+) patients intervened by ambulances teams according to medical mask availability is analysed (As shown Table 7). The number of patients with medical mask on is 2200 and the number of patients without mask is 161. When the patient wear mask, the risk level of HCWs is decreasing. If the patient no mask available, 16.10% of HCWs are under the medium risk and 35.40% under low risk. The relation between using mask on patient and risk level of HCWs is statistically significant (p < 0.001). The additional strategies and decision making mechanisms which are used by HCWs during emergencies could not be searched and examined due to time restriction and over workload during outbreak.

**Table 7:** The Medical Mask Use for Patients by Risk Level of HCWs (N: 2361)

| The using of medical mask on patients | No risk | | Low risk | | Medium risk | | Total | | $\chi^2$<br>p |
| --- | --- | --- | --- | --- | --- | --- | --- | --- | --- |
|  | N | % | N | % | N | % | N | % | *272.072<br>P < 0.001 |
| Mask available | 2003 | 91.00 | 156 | 7.10 | 26 | 1.90 | 2200 | 100 |  |
| No mask available | 78 | 48.40 | 57 | 35.40 | 41 | 16.10 | 161 | 100 |  |
| Total | 2081 | 88.10 | 213 | 9.00 | 67 | 2.80 | 2361 | 100 |  |

\* *Kendall's tau-c*

Many HCWs are infected during COVID-19 outbreak (49, 50). During this study conducted, there are only 2 HCWs infected with COVID-19 diagnosed by CT screening. One of them from 112 call centre, the other one from ambulance station and both of them are evaluated with no risk available. Since no risk or exposure to COVID-19 (+) staff and/or patient during their shifts, they could have been infected when out of duty. No other staff has been found infected by COVID-19 from their shift. The follow-up procedures of infected HCWs have been finalized, both of them cured.

## 4. CONCLUSION AND RECOMMENDATIONS

The level of PPE use, other measures against COVID-19 and awareness of HCWs are considered to be effective to achieve the results of this study. During the study conducted between March 14th, 2020 – May 31st, 2020; Antalya Ambulance teams intervened to 46% of whole COVID-19 confirmed cases in Antalya. The rest of confirmed patients in Antalya, have directly applied to hospital, family physician, and other healthcare facilities or found by filiation (51) method. The ambulance teams reached to 5344 COVID-19 possible cases and 787 confirmed cases. The majority of confirmed cases were male (62%) and over the age of 65 (42%). Most of the confirmed cases are transferred to healthcare facilities (75%).

The total 2361 HCWs were taken to risk analysis and then followed-up procedures. The 67 of them were found under the medium risk, 213 under the low risk and 2081 of them evaluated with no risk. Level-4 PPE which is consisted of N95/FPP – Goggle / face protection – Gloves –Apron / coverall, mostly used (84.50%) for confirmed cases by HCWs. The level of PPE usage showed negative correlation compare to risk level. When the level of PPE used increased, risk level of HCWs was decreased. The risk level of HCWs was also decreased, if medical mask used for patients. The 16.10% of HCWs were under medium risk, and 35.40% of them under low risk, when patients wear no mask on.

There were only 2 HCWs diagnosed with COVID-19: one of them from 112 call centre, and other one is from one of the ambulance station. Both of them were not exposed to COVID-19 (+) staff or patient during their shifts. COVID-19 exposure might had happened while they are off work. The follow-ups of HCWs have been finalized, and both of them cured.

Ongoing infection prevention and control studies in Antalya 112 Emergency Ambulance Services, have been effective in reducing the risk of HCWs exposure to COVID-19 and management of outbreak. Providing the supply of the PPE needed in time and without interruption, work planning, staff training program implementation, alternative scenarios planning for unexpected situations, staff participation in decision making process, were effective in managing the outbreak.

Furthermore, alternative plans and strategies specific to countries, regions and type of healthcare facilities; should be developed and implemented by following global practices and innovations. The future studies about easy-to-use, high-tech PPE with maximum protection are recommended.

## Data Availability

In this study, data (COVID-19 confirmed case information, level of PPE usage of ambulance teams, and risk analysis results of follow-up measures of HCWs) were obtained from Directorate of Antalya Emergency Ambulance Services. The level of risk analysis results, follow-up measures of HCWs are obtain by the COVID-19 response team and ASOS records (Emergency Health Information Systems).

